# Gene expression differences associated with alcohol use disorder in human brain

**DOI:** 10.1101/2024.01.20.24301386

**Authors:** Caryn Willis, Julie D. White, Melyssa S. Minto, Bryan C. Quach, Shizhong Han, Ran Tao, Joo Heon Shin, Amy Deep-Soboslay, Thomas M. Hyde, R. Dayne Mayfield, Bradley T. Webb, Eric O. Johnson, Joel E. Kleinman, Laura J. Bierut, Dana B. Hancock

## Abstract

Excessive alcohol consumption is a leading cause of preventable death worldwide. Neurobiological mechanisms associated with alcohol use disorder (AUD) remain poorly understood. To further understand differential gene expression (DGE) associated with AUD, we compared deceased individuals with and without AUD across two human brain regions, nucleus accumbens (NAc) and dorsolateral prefrontal cortex (DLPFC). Bulk RNA-seq data in both NAc and DLPFC from human postmortem brains (N ≥ 50 with AUD and ≥ 46 non-AUD) were analyzed for DGE using negative binomial regression adjusting for technical and biological covariates. The region-level results were meta-analyzed with a previously published, independent dataset (N_NAc_= 28 AUD, 29 non-AUD; N_PFC_= 66 AUD, 77 non-AUD). We further utilized these data to test for heritability enrichment of AUD-related phenotypes, gene co-expression networks, gene ontology enrichment, and drug repurposing. We identified 176 differentially expressed genes (DEGs; 12 in both regions, 78 only in NAc, 86 only in DLPFC) for AUD in our new dataset. By meta-analyzing with published data, we identified 476 DEGs (25 in both regions, 29 only in NAc, 422 only in PFC). Of these DEGs, we found 17 genes that were significant when looked up in GWAS of problematic alcohol use or drinks per week. Gene co-expression analysis showed both concordant and unique gene networks across brain regions. We also identified 29 and 436 drug compounds that target DEGs from our meta-analysis in NAc and DLPFC, respectively. This study identified robust AUD-associated DEGs, providing novel neurobiological insights into AUD and highlighting genes targeted by known drug compounds, generating opportunity for drug repurposing to treat AUD.

## Introduction

Alcohol use disorder (AUD) affected approximately 28.6 million adults in 2021 in the United States^1^, and there are 3 million deaths per year caused by harmful use of alcohol world wide^2^. AUD has 50-60% heritability^3^, and hundreds of genome-wide significant variants have been identified for alcohol dependence^4^, problematic alcohol use^5–7^, and/or consumption^7–9^ (e.g., drinks per week). As a more complete picture is emerging for specific genetic variants underlying AUD and related phenotypes, the gene regulatory landscape associated with AUD remains largely unknown. Filling this critical gap will define potentially new neurobiological mechanisms associated with AUD and assist in the identification of possible new drug targets to treat AUD. By focusing on gene expression changes associated with AUD in human brain, this study identifies regulatory differences that may be driven by predisposing genetic variation or may be consequences of the alcohol exposure; both improve our understanding of the neurobiological mechanisms that relate to AUD.

Our study compares data from AUD cases and non-AUD controls in two key brain regions involved in the addiction cycle: the nucleus accumbens (NAc), implicated in the binge/intoxication stage, and the dorsolateral prefrontal cortex (DLPFC), implicated in the preoccupation/anticipation stage^10^. Both brain regions are linked to reward pathways as components of the dopaminergic mesolimbic system^11^. Exposure to alcohol increases dopamine levels via effects on dopaminergic neurons that originate in the midbrain and project into forebrain regions with NAc and prefrontal cortex (PFC) being most relevant to reward processes and addiction^12,13^. Disruption of the dopamine system in the NAc has been described as lying at the core of addiction^14^. Dopamine release into the NAc is regulated by the PFC^15^ and PFC dysfunction is associated with impulsivity, compromised executive function, and increased engagement in risky behavior^16^. Thus, the NAc and PFC are distinct, yet interrelated brain regions with functions highly relevant to the molecular mechanisms of AUD.

A few prior studies of AUD-related bulk RNA-seq gene expression in human brain have been reported, providing initial evidence for differential gene expression (DGE)^17,18,19,20,21^. However, none of these studies assessed results across independent datasets. Kapoor *et al*. conducted the largest prior study (N=138) with RNA-seq in PFC and identified 129 genes that showed significant altered expression (FDR < 0.05) between the 65 AUD cases and 73 non-AUD controls^17^. A smaller subset with RNA-seq in NAc (N=30 AUD, 30 non-AUD) showed 14 genes with significant altered expression^19^. Zillich *et al*. reported another dataset (N=48 AUD, 51 non-AUD) with RNA-seq in three other brain regions and found significant evidence for DGE related to AUD in caudate nucleus (CN; 49 genes FDR < 0.05) and putamen (PUT; 1 gene FDR < 0.05) but no significant evidence in the ventral striatum (VS)^18^.

In this study, we report on a new dataset of 96 RNA-seq samples from NAc and 98 from DLPFC and meta-analyze results with uniform reprocessing of the Kapoor *et al*. dataset in NAc and PFC to increase sample size and statistical power to identify genes with robust evidence for DGE. We further evaluated overlap of differentially expressed genes (DEGs) between our study and Zillich *et al*. to compare DGE across brain regions, integrated DEGs with GWAS results to infer genetically driven DGE, and conducted gene ontology enrichment and gene co-expression analyses to explore potential mechanisms of DGE in AUD. We also looked up our AUD-associated DEGs in drug repurposing databases to identify pharmacotherapies that might target AUD.

## Materials & Methods

A workflow overview of the datasets and analyses can be found in Figure 1.

**Figure 1.**
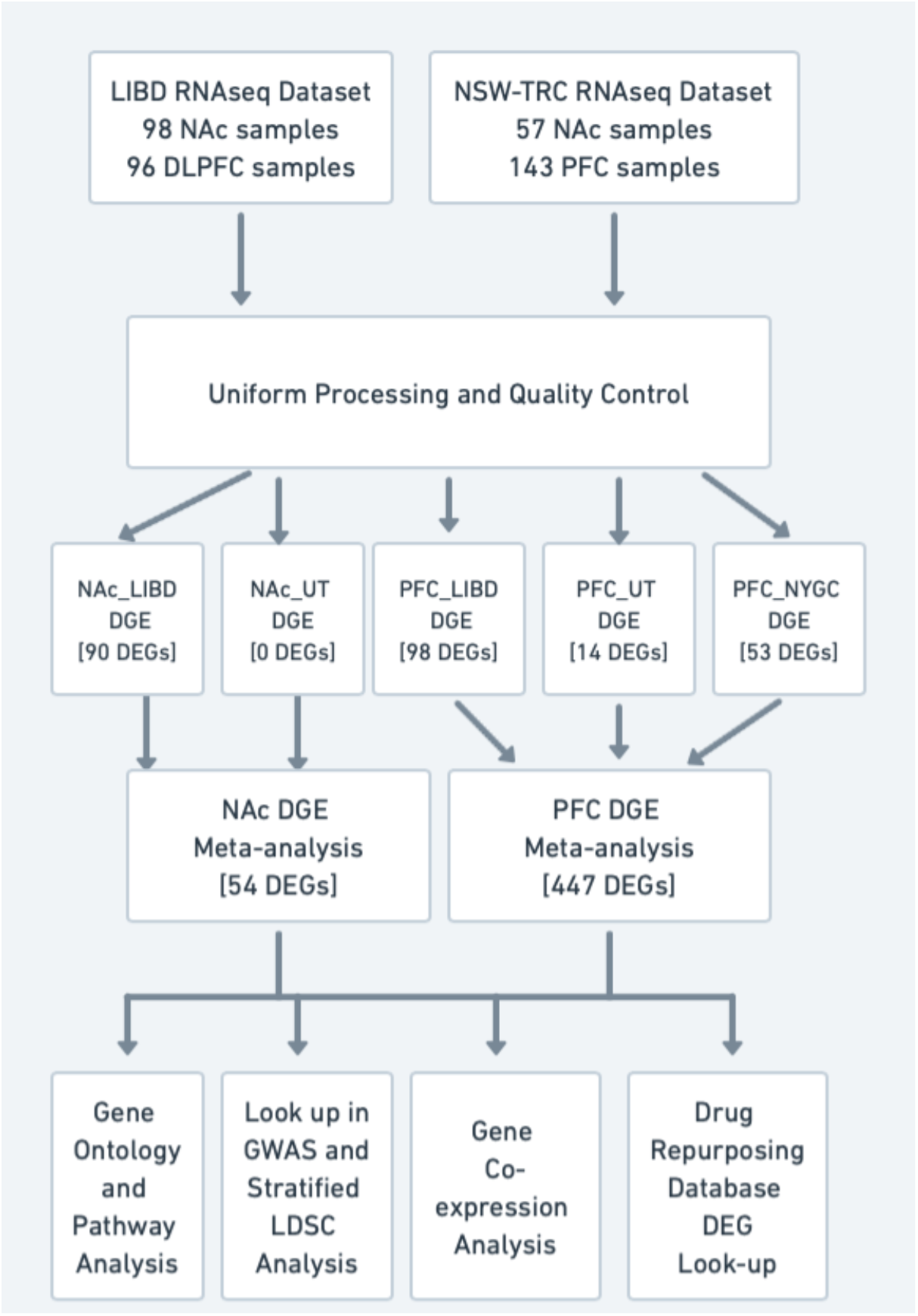
Overview of analysis workflow.

### AUD cases and controls among human postmortem NAc and PFC samples

Postmortem human DLPFC (BA 46/9) and NAc tissues were obtained at autopsy from 122 subjects (61 AUD, 61 non-AUD) as part of the Lieber Institute for Brain Development (LIBD) Human Brain Repository^22^. AUD cases and controls were defined based on two or more lifetime DSM-5 symptoms within a 12-month period. Non-AUD controls had no lifetime history of DSM-5 AUD symptoms and postmortem ethanol toxicology of less than 0.06 g/dL. Decedents with major depressive disorder (MDD) were defined as those with a lifetime history of five or more DSM-5 MDD symptoms persisting for two weeks or longer. All details regarding these samples and phenotype data were provided in White *et al*.^23^ RNA was extracted using LIBD’s existing protocol^24,25^. Illumina TruSeq Total RNA Stranded RiboZero Gold (Illumina Inc, San Diego, CA) was used for library prep. These samples will be referred to as the NAc_LIBD and PFC_LIBD datasets.

The Kapoor *et al*. samples were obtained from the New South Wales Tissue Resource Center (NSW-TRC) and sequenced in two batches, one at the University of Texas at Austin (UT Austin) and the other at the New York Genome Center (NYGC). From both batches, fastq files for 143 prefrontal cortex (BA8) samples and 58 NAc samples were transferred from the University of Texas at Austin (Table 1). The raw data is publicly available on NCBI (PRJNA551775, PRJNA530758, PRJNA781630). Sequencing information was detailed in Kapoor *et al*^17^. The samples that were processed at UT Austin will be referred to as NAc_UT and PFC_UT. The samples processed at the New York Genome Center (NYGC) will be referred to as PFC_NYGC.

**Table 1.**
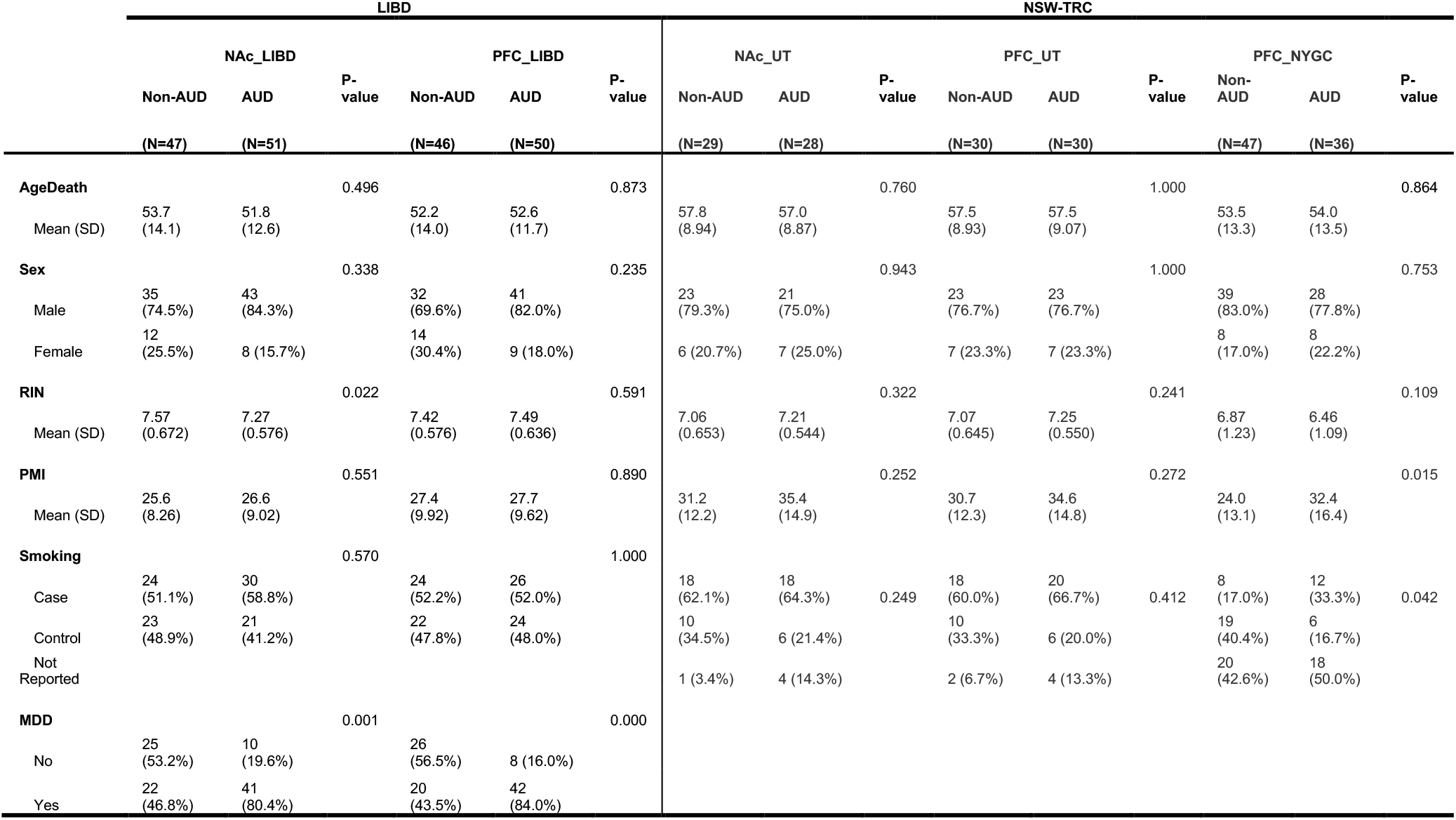
Demographic information of samples in each dataset.

Given that BA 46/9 (LIBD) and BA8 (NSW-TRC) are cytoarchitecturally adjacent and involved in similar brain functions, we chose to meta-analyze data from the two to maximize power. When discussing results specific to the LIBD samples, we use DLPFC to distinguish, when discussing results specific to the NSW-TRC samples, we use PFC, and when discussing meta-analysis results, we use PFC.

### Bulk RNA-seq data processing, quality control (QC)

For all datasets, reads were trimmed and filtered using Trimmomatic^26^ and transcripts were quantified using the GENCODE v40 (GRCh38) transcriptome with Salmon (v 1.1.0)^27^. Gene-level quantification was done with the tximport R package (v1.14.2)^28^. QC metrics were calculated using MultiQC v1.7^29^. Samples were filtered using the following QC metrics: effective sequencing depth >10 million reads, post-Trimmomatic retained reads percentage > 60%, mean read GC content between 35% and 65%, Salmon mapping percentage > 30%, gene mapping percentage (combined intronic and exonic mapping) > 80%, intergenic/genic mapping ratio < 0.9, Shannon index > the Shannon index IQR-based cutoff or transcriptome mapping percentage > 50%, mitochondrial mapping percentage < 10%, ribosomal RNA mapping percentage < 1%, and RIN > 5. After QC filtering, 98 samples (51 AUD, 47 Non-AUD) remained for NAc_LIBD and 96 (50 AUD, 46 Non-AUD) samples for PFC_LIBD (Table 1). Because the PFC samples from Kapoor *et al*. were sequenced at two different locations (UT Austin and NYGC) and had different sequencing depths, samples from each location were kept separate for analysis. After QC filtering, 57 (28 AUD, 29 non-AUD) NAc_UT samples remained, and all 143 (66 AUD, 77 Non-AUD) PFC samples remained (60 from PFC_UT, 83 from PFC_NYGC).

### Cell type deconvolution

To estimate cell-type proportions for each dataset, the Bisque R package (v1.0.5)^30^ was used along with single-cell references^31^ for both NAc and DLPFC. Because both the NAc single cell reference and NAc_LIBD use data from LIBD, one sample coincidently overlapped. This sample was indicated to Bisque as overlapping. Proportions for macrophage, microglia, oligodendrocyte progenitor cells (OPCs), astrocytes, GABAergic neurons, oligodendrocytes, and medium spiny neurons (MSNs) were estimated for NAc. Proportions for mural cells, macrophage, T-cell, microglia, OPCs, astrocytes, GABAergic neurons, excitatory neurons, and oligodendrocytes were estimated for PFC. The differences in cell types by region were due to cell type differences in the reference datasets. Linear regression was used to test the association between cell-type and AUD status, accounting for MDD, smoking, age, sex, PMI, and RNA Integrity Number (RIN) for NAc_LIBD and PFC_LIBD and age, sex, postmortem interval (PMI), and RIN for the NAc_UT, PFC_UT, and PFC_NYGC. No cell-type proportion differences were significantly associated with AUD in any of the datasets (Supplemental Table 1). This motivated our decision to not include cell type proportion estimates in our DGE models.

### Differential gene expression analysis

Genes were filtered for low expression using a cutoff of 10 counts in at least the number of samples that make up the smaller AUD status group for a given dataset. Surrogate variables (SVs) were calculated using the sva R package (v3.42.0)^32^ to serve as proxies for known covariates and unmeasured technical and biological confounds. Percent variance explained was used for model selection. Each model had an r^2^ value greater than 0.6 including model covariates other than AUD. We used the DESeq2 R package (v1.34.0)^33^ to test for DGE using a generalized linear model with gene expression as the outcome variable. The following models were used for DGE analyses:

NAc_UT: expression ∼ AUD + SV1 + … + SV9

NAc_LIBD: expression ∼ AUD + MDD + smoking status + SV1 + … + SV12

PFC_UT: expression ∼ AUD + SV1 + … + SV10

PFC_NYGC: expression ∼ AUD + SV1 + … + SV10

PFC_LIBD: expression ∼ AUD + MDD + smoking status + SV1 + … + SV11

DEGs were assessed for potential sample outlier effects and removed based on Cook’s distance using a threshold of over the 99^th^ percentile of the F-distribution. A Benjamini-Hochberg false discovery rate (FDR) of 0.05 was used to declare statistically significant DGEs.

### Differential gene expression meta-analyses

Meta-analyses were performed for each brain region using the weighted Fisher’s method in the metaPro R package (v1.5.5), which incorporates sample sizes of contributing datasets. Only genes tested for DGE across all datasets were considered for meta-analysis. An FDR of 0.05 was used to declare statistical significance. Genes that had discordant fold changes between datasets were removed after significance calling for consistency and robustness.

### Lookup of meta-analysis DEGs in summary stats of other brain regions

To compare our results across brain regions, we looked up our meta-analysis DEGs in the summary statistics of the Zillich *et al*.^18^ CN, VS, and PUT DGE analyses associated with AUD. A Bonferroni-corrected p value was used to declare significance, correcting for the number of meta-analysis genes looked up in each of the brain region summary statistics (p < 0.05/48 for NAc and p < 0.05/401 for PFC).

### Gene ontology and pathway analysis

ToppFun from the ToppGene Suite^34^ was used to detect functional enrichment of genes implicated in the DGE meta-analysis using the following GO databases: Molecular Function, Biological Process, Cellular Component, and KEGG Pathway. FDR < 0.05 was used to declare a statistically significant term.

### Lookup of DEGs in GWAS and Stratified Linkage Disequilibrium Score Regression (LDSC)

We performed a lookup of DEGs in gene level summary statistics for problematic alcohol use^5^ (gene level using H-MAGMA^35,36^) and drinks per week^9^ (gene level using MAGMA^37^) using Bonferroni corrected significance p-value < 0.05, correcting for the number of our meta-analysis DEGs that appear in the GWAS gene level summary statistics.

Partitioned heritability estimates and tests for enrichment of genetic loci associated with AUD-related phenotypes (alcohol consumption as measured by drinks per week ^9^ and alcohol dependence^4^) constrained to DGE loci were conducted using stratified LDSC (v1.0.1)^38,39^. An annotation window of 100kb from start and 100kb from end of the meta-analysis significant genes was tested. A Bonferroni corrected (corrected for 2 phenotypes tested) p-value threshold of 0.025 was used to declare significance.

### Gene co-expression analysis

The weighted gene co-expression network analysis (WGCNA) R package (v1.71)^40^ was used to construct gene networks, with hierarchical clustering and dynamic tree-cutting to define modules for each dataset. The PFC_UT and PFC_NYGC were combined for this analysis to increase sample size. Soft power was set for each brain region and dataset where the scale-free topology fit index reached 0.90. Additional settings used were minimum module size = 100, cutting height = 0.99, deepSplit = TRUE, and pamStage = FALSE. Modules were merged based on an eigengene (*i*.*e*., top gene expression principal component of genes in a module) correlation threshold of 0.75. Covariates were accounted for using the removeBatchEffect function from the limma R package (v3.50.0)^41^. For NAc_LIBD and PFC_LIBD, MDD, smoking, sex, and age were used as covariates. For NAc_UT, PFC_UT and PFC_NYGC, batch and RIN were used as covariates, as done in Kapoor *et al*. Since the soft threshold step failed to converge for the NAc_UT dataset, that dataset was excluded from this analysis. We looked at module similarity across brain regions within the LIBD dataset by calculating the percentage of overlap between modules, defined as the number of overlapping genes divided by size of the smaller of the two modules. We tested for association between module eigengenes and AUD status using an ANOVA test.

### Meta-analysis DEG look-up in drug repurposing databases

To identify the potential druggability of genes with AUD-related DGE, we used a drug repurposing tool^42^, which leverages four different drug repurposing databases (Pharos, Open Targets, Therapeutic Target Database, and DrugBank) and a ranking system based on association statistics to provide a ranked list of drug compounds that target genes of interest. The lists of meta-analysis DEGs ranked by FDR-corrected p-values were used for the analysis. For NAc, all DEGs were used for the analysis. Given the large number of AUD-associated genes with differential expression in PFC, to focus on the most significant results, the top 100 DEGs for PFC were used for the analysis. To filter the results for drugs that have specific targets, the top 10th percentile of gene target ratio was selected to focus more on targeted therapies. The gene target ratio is calculated as the number of genes in our gene set that a given drug targets divided by the total number of genes that the drug targets.

## Results

### Differential gene expression results for each dataset

Each independent dataset was tested separately for DGE to account for batch effects. DGE analysis of 51 AUD and 47 non-AUD samples from the NAc_LIBD dataset resulted in 90 DEGs at FDR < 0.05. DGE analysis of 50 AUD and 46 non-AUD samples for PFC_LIBD dataset resulted in 98 DEGs at FDR < 0.05 (Figure 2A, Supplemental Table 2). Twelve genes overlapped between the significant results from NAc_LIBD and PFC_LIBD.

**Figure 2.**
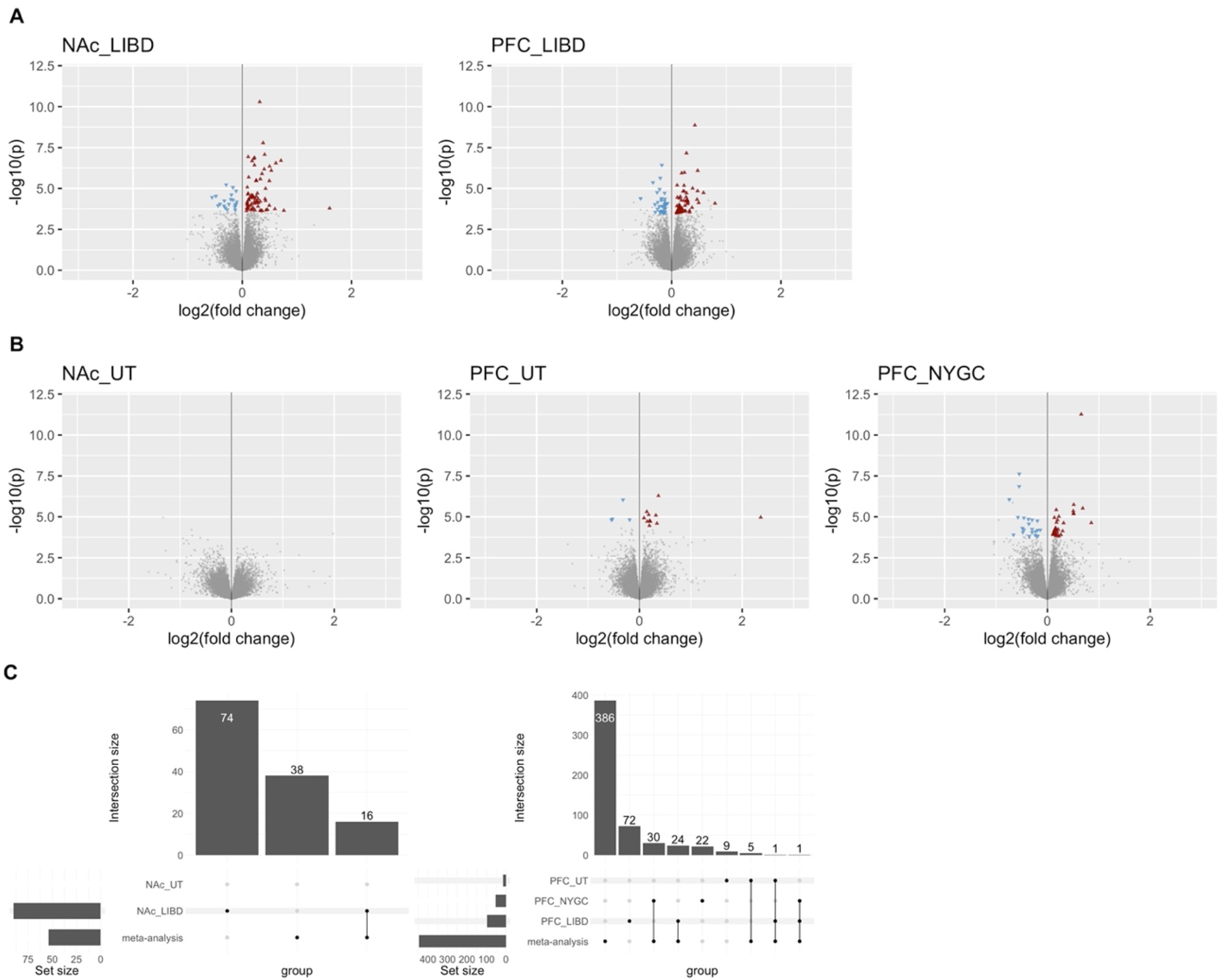
(A) Volcano plots of the differentially expressed genes (FDR < 0.05) by AUD status in the LIBD datasets. Blue triangles represent significant downregulated genes and red triangles represent significantly upregulated genes. Genes not significantly differentially expressed are colored grey. (B) Volcano plots of the differentially expressed genes (FDR < 0.05) by AUD status in the NSW-TRC datasets. Blue triangles represent significant downregulated genes and red triangles represent significantly upregulated genes. Genes not significantly differentially expressed are colored grey. (C) Upset plots displaying the gene overlap count between the individual datasets DEGs and meta-analysis DEGs by brain region.

Uniform processing and QC (matching the processing and QC of NAc_LIBD and PFC_LIBD) of the NAc_UT resulted in 28 AUD and 29 non-AUD samples after QC filtering. For the PFC_UT 30 AUD and 30 non-AUD samples were available for analysis. For PFC_NYGC, 36 AUD and 47 non-AUD samples were available for analysis. DGE analysis resulted in no DEGs in the NAc_UT dataset, and 14 and 53 DEGs (FDR < 0.05) with no overlapping genes, in PFC_UT and PFC_NYGC, respectively (Figure 2B).

### DGE meta-analyses across datasets

Meta-analysis across the NAc samples resulted in 54 DEGs (33 upregulated and 21 downregulated) at FDR < 0.05 with the top 5 DEGs genes being *ODC1, ZNF844, ARRDC3, FAM225A*, and *GUSBP11*. Meta-analysis across the PFC samples resulted in 447 DEGs (295 upregulated and 152 downregulated) at FDR < 0.05 with the top 5 DEGs genes being *TXNIP, ODC1, HMGN2, SLC16A9*, and *SLC16A6* (Supplemental Table 3). Only genes with same direction of effect were declared significant. Twenty-five DEGs overlapped between FDR significant results for NAc and PFC. Overlap between individual studies and the meta-analyses by brain region are shown in Figure 2C.

### Lookup of meta-analysis DEGs in summary statistics of other brain regions

After Bonferroni correction for the NAc meta-analysis genes, *TTLL4* was the only gene significantly associated with AUD in CN, while no NAc meta-analysis genes were associated with AUD in VS and PUT. After Bonferroni correction, no PFC meta-analysis significant genes were significantly associated with AUD in CN, VS, or PUT. The lack of overlap between NAc and PFC results with CN, VS, and PUT results suggests brain region specific-DGE associated with AUD.

### Gene ontology and pathway analyses

To assess the functions and pathways of genes implicated in our DGE meta-analysis, we performed ToppGene enrichment for genes in each brain region then used semantic similarity to cluster the results. ToppGene enrichment analysis for DEGs from the NAc meta-analysis resulted in enrichment of 3 GO Molecular Function terms at FDR < 0.05: choline kinase activity, glucosyltransferase activity, and ethanolamine kinase activity. The enrichment analysis for DEGs from the PFC meta-analysis resulted in no significant enrichment (Supplementary Table 4).

### GWAS overlap for genetically driven gene expression

To investigate the connection between genetic variation and DGE associated with AUD, we performed a lookup of our meta-analysis significant genes with AUD-related GWAS summary statistics and used stratified LD score regression to test enrichment of GWAS signals in meta-analysis DEGs. For the problematic alcohol use^5^ GWAS, 50 NAc DEGs were tested, and one gene (*UGGT2*) were significantly associated after Bonferroni correction (p < 0.05/50); 424 PFC DEGs were tested, and six genes (*TCTA, TSPAN5, TLR6, EMX2OS, CA11*, and *SHISA5*) were significant (p < 0.05/424). For the drinks per week (DPW) GWAS^9^, 41 NAc DEGs were tested, and two genes (*GBA2* and *GRM5*) were significant (p < 0.05/41); 379 PFC DEGs were tested, and nine genes (*HDAC7, HIPK3, ACVR2B, CDH18, TSPAN5, TTLL6, OLFML2A, SIRPA*, and *ATXN1L*) were significant (p < 0.05/379).

Testing the meta-analysis DEGs for enrichment in GWAS with fully available summary statistics (DPW^9^ and alcohol dependence^4^) using stratified LDSC resulted in enrichment (Bonferroni correction for two traits) for DPW in the PFC (Enrichment = 1.46, p = 0.0131) (Supplementary Table 5). Both the lookup in gene-level GWAS and the stratified LDSC analysis of SNP-level GWAS suggest there may be a genetic component to some of the AUD-associated DGE.

### Gene co-expression analysis

WGCNA was used to explore the co-expression of genes within the datasets and test for module association with AUD. WGCNA generated 15 modules for the PFC_LIBD dataset, 14 modules for the NAc_LIBD dataset, and 22 modules for the PFC_UT and PFC_NYGC combined dataset (Supplementary Table 6). After FDR correction, no modules were associated with AUD. Module consistency across brain regions was tested by calculating the percentage of overlapping genes between modules. Comparing NAc_LIBD and PFC_LIBD modules, each module overlapped with at least one other module with a minimum of 39.2% and maximum of 97.8% sharing (Supplementary Figure 2), which supports prior evidence that there is both shared and region-specific co-expression across brain regions^43^.

### DEG look-up in drug repurposing databases

Using the Drug Repurposing Database tool^42^ and our meta-analysis DEGs, 11 of the 54 genes with AUD-associated DGE in NAc were targeted by 29 drug compounds (Supplemental Table 7). 64 of the top 100 genes with AUD-associated DGE in DLPFC were targeted by 436 drug compounds. To focus more on targeted therapies, the 436 compounds were then subset to 67 compounds that have the highest 10% of prioritized genes to all genes targeted ratio (ratio ≥ 0.33). This step was not conducted for the NAc results due to the much smaller numbers of DEGs and compounds.

## Discussion

This study reports many newly identified DEGs, providing neurobiological insights into gene expression signatures of AUD that are shared across or specific to key brain regions for addiction. For NAc, there were no DEGs in the NAc_UT dataset, when analyzed alone, but when uniformly processed and meta-analyzed with our NAc_LIBD dataset, we identified 54 DEGs associated with AUD. By uniformly processing and meta-analyzing our new PFC_LIBD dataset with the previously published PFC_UT and PFC_NYGC datasets, we were able to increase the number of DEGs from 129 that Kapoor *et al*. reported to 447 DEGs for PFC. Overall, 25 genes were differentially expressed with AUD in both NAc and PFC. Unlike prior studies, independent datasets were combined via meta-analysis, enabling us to model the complexities of each dataset and take advantage of the combined sample size for more statistical power to detect AUD-associated DGE.

Of the 54 significantly expressed genes in the NAc meta-analysis, a lookup in the Zillich *et al*. summary statistics of VS, CN, and PUT regions resulted in only one significant gene (*TTLL4*) that overlapped between our NAc results and the CN results. These results provide initial evidence for cross-region DGE in relation to AUD, though many genes appear to have tissue-specific DGE. We also observed both shared brain region expression and region specificity through the cross-region WGCNA module analysis.

The overlap of several of our meta-analysis DEGs in alcohol behavior GWAS suggests that at least some of the AUD-associated DGE may be genetically driven. By conducting a stratified LDSC analysis, we tested for enrichment of alcohol behavior-associated genetic loci in and around DEGs for AUD. Significant enrichment between the drinks per week summary statistics and PFC DEGs further supports a genetic component to the DEGs.

Many of the top DEGs and DEGs that overlapped with GWAS results were previously reported to play a role in AUD or other psychiatric disorders. *ODC1*, which was significant in both NAc and PFC meta-analyses and associated with cell proliferation of neural progenitor cells, was reported to be downregulated in psychiatric phenotypes by loss of function variants^44^. *TXNIP*, a top DEG in the PFC, was upregulated in mice PFC in schizophrenia-like states^45^ and implicated in astrocytic glucose hypometabolism in depressive state rats^46^.

Differential expression of *HMGN2*, a top DEG in the PFC, was reported for schizophrenia-associated DGE in peripheral blood mononuclear cells ^47^. *SLC16*, a top DEG in the PFC, with many other solute carrier transporters having reported roles in neuropsychiatric disorders, such as *SLC6* in depression and post-traumatic stress disorder^48^. *TSPAN5*, a DEG in the PFC and significant in both PAU and DPW GWAS, is downregulated by acamprosate^49^, one of only three FDR-approved treatments for AUD. *TLR6*, a Toll-Like Receptor (TLR), was a DEG in the PFC and significantly associated in problematic alcohol use GWAS. Dysregulation of TLRs, which are directly involved in the regulation of inflammatory reactions, has been reported in AUD due to an inflammatory response to excessive alcohol consumption^50^. *CDH18*, a DEG in the PFC and significantly associated in DPW GWAS, and other cadherin pathway genes have been associated with schizophrenia, bipolar disorder and MDD through GWAS^51^. These converging associations could indicate a shared genetic, and ultimately transcriptomic, risk for AUD and some psychiatric disease, a hypothesis supported by shared genetic correlations^52^. While several of our DEGs have known associations with AUD and other psychiatric disorders, many of the DEGs are novel to the AUD phenotype and their roles in AUD should be investigated further.

By integrating our DGE results with drug repurposing databases, we identified many drug compounds that target the DEGs in both NAc and DLPFC. Some of the listed compounds are already used for AUD treatment, such as clomethiazole which targets the GABA receptors and is used for alcohol withdrawal treatment^53^ and acamprosate, which targets *GRM5* and is used for treatment of alcohol dependence. *GRM5* also overlapped between our NAc DGE results and the drinks per week GWAS. *GRM5* is also targeted by cinacalcet, rufinamide, and glutamic acid, which could be considered as candidate drugs to treat AUD with support from both GWAS and differential expression. Another gene with overlapping support from both DGE and GWAS was *GBA2*, which is targeted by miglustat and used for treatment of Gaucher’s Disease^54^. Though we identified some compounds already used for AUD treatment, lending confidence to our approach, most of the compounds that we identified are used for purposes unrelated to AUD and merit further investigation as potential novel therapeutics for AUD.

This study comes with limitations. The data are limited to European ancestry, so results merit testing in other datasets of diverse ancestries to assess their generalizability. Though this is the largest combined sample size of human brain samples in NAc and PFC to date (N up to 239), even larger sample sizes across diverse populations and brain regions that play a role in the addiction cycle are needed. This study is also based around lifetime history of AUD, so some gene expression could have returned to control levels over time if drinking was reduced or eliminated prior to death, biasing the results for those genes towards the null. No estimated cell type proportions in any of the datasets were significantly associated with AUD, though this analysis was limited by the small sample sizes available for single-nuclei RNA-seq datasets.

Because estimation is fully based on the reference dataset, having a small reference dataset can lead to bias. There was also only overlap of one sample with the reference data which affects the estimation. Since the estimation only reflects the proportions seen in the single-nuclei reference dataset, using a larger sample size dataset in the future and a larger overlap with bulk samples will help improve this analysis.

This work is the first meta-analysis of DGE of AUD in NAc and DLPFC human brain regions. Meta analyzing DGE results from two different studies using uniform processing and analysis helped identify more robust results than from one study alone. Many identified top DEGs are known to be involved in AUD and other psychiatric disorders, while others are novel. Overlap of several DEGs with alcohol use-related GWAS and enrichment in GWAS of drinks per week, suggests that these DGE findings may have a genetic component. By using a drug repurposing tool, we were able to identify pharmacotherapies that have already been used for AUD as well as many novel candidate pharmacotherapies for AUD.

## Acknowledgements

The authors would like to gratefully acknowledge the generosity of the families of the decedents, who donated the brain tissue and used in this study. We are also thankful for the many colleagues whose efforts have led to the donation and curation of the postmortem tissue that makes this study possible.

## Disclosures

### Data availability

Bulk RNAseq data generated from this study has been deposited on GEO (GSE253155) and summary statistics for analyses are provided as supplement tables.

### Funding

This work was supported by the National Institute on Alcohol Abuse and Alcoholism R01 AA027049 (mPI: Hancock & Bierut).

### Conflict of Interest

The following authors declare no conflict of interest: CDW, JDW, MSM, BCQ, SH, RT, JHS, AD-S, TMH, RDM, BTW, EOJ, and DBH.

JEK is a member of a drug monitoring committee for an antipsychotic drug trial for Merck. LBJ is listed as an inventor on U.S. Patent 8,080,371, ‘Markers for Addiction’ covering the use of certain SNPs in determining the diagnosis, prognosis, and treatment of addiction.

## Notes

### Author Declarations

The IRB committee of RTI International waived ethical approval for this work.

