## Supplementary figures and images for "Gene expression differences associated with alcohol use disorder in human brain"

### Supplemental Figure 1

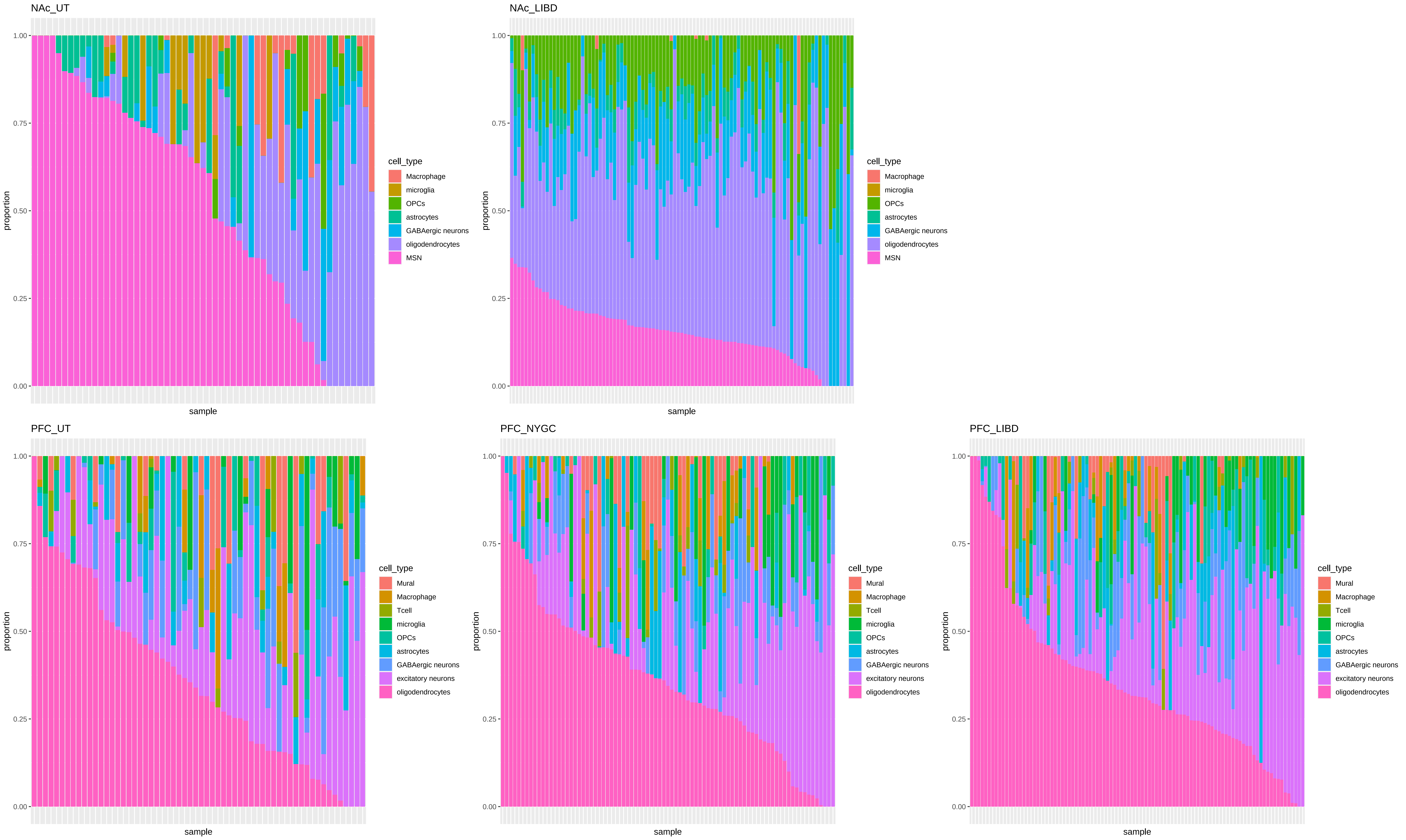

### Supplemental Figure 2

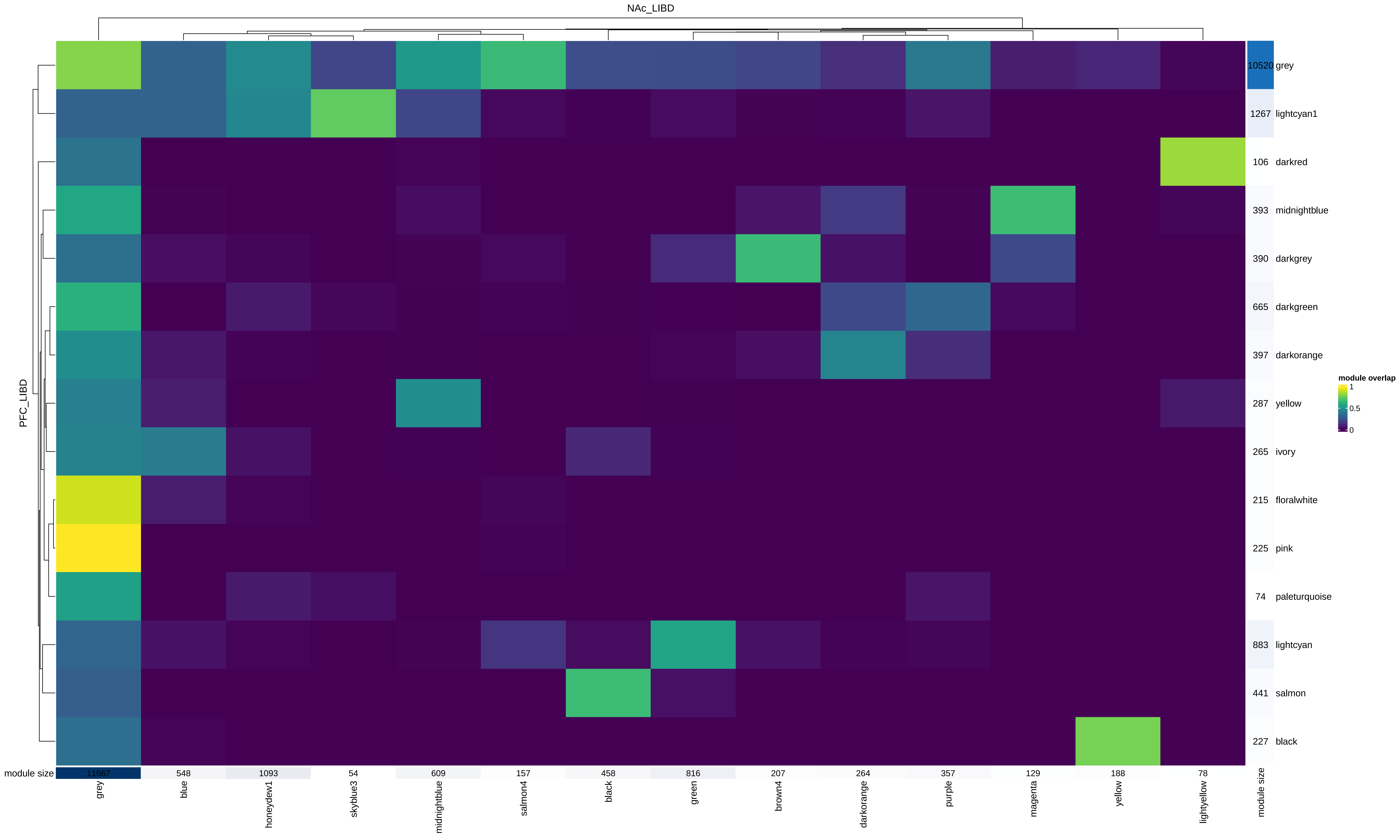
